# Can artificial intelligence diagnose seizures based on patients’ descriptions? A study of GPT-4

**DOI:** 10.1101/2024.10.07.24314526

**Authors:** Joseph Ford, Nathan Pevy, Richard Grunewald, Steve Howell, Markus Reuber

**Author notes:** Corresponding author: Joseph Ford.

## Abstract

**Introduction:** Generalist large language models (LLMs) have shown diagnostic potential in various medical contexts. However, there has been little work on this topic in relation to epilepsy. This paper aims to test the performance of an LLM (OpenAI’s GPT-4) on the differential diagnosis of epileptic and functional/dissociative seizures (FDS) based on patients’ descriptions.

**Methods:** GPT-4 was asked to diagnose 41 cases of epilepsy (n=16) or FDS (n=25) based on transcripts of patients describing their symptoms. It was first asked to perform this task without being given any additional training examples (‘zero-shot’) before being asked to perform it having been given one, two, and three examples of each condition (one-, two, and three-shot). As a benchmark, three experienced neurologists were also asked to perform this task without access to any additional clinical information.

**Results:** In the zero-shot condition, GPT-4’s average balanced accuracy was 57% (κ: .15). Balanced accuracy improved in the one-shot condition (64%, κ: .27), though did not improve any further in the two-shot (62%, κ: .24) or three-shot (62%, κ: .23) conditions. Performance in all four conditions was worse than the average balanced accuracy of the experienced neurologists (71%, κ: .41).

**Significance:** Although its ‘raw’ performance was poor, GPT-4 showed noticeable improvement having been given just one example of a patient describing epilepsy and FDS. Giving two and three examples did not further improve performance, but more elaborate approaches (e.g. more refined prompt engineering, fine-tuning, or retrieval augmented generation) could unlock the full diagnostic potential of LLMs.

## 1. Introduction

While the medical potential of artificial intelligence (AI) has been discussed and studied for decades,^1^ the public release of OpenAI’s ChatGPT in late 2022 (and the subsequent release of other, similar assistants) has driven a new wave of interest in the topic. This interest extends to the field of epilepsy, where the potential of AI is now clearly recognised.^2^

There are several reasons for the recent excitement about AI. First, earlier research tended to focus on specialist AI systems that had been trained to perform well on limited, domain-specific tasks. In contrast, ChatGPT and other such assistants are based on generalist large language models (LLMs), trained to identify patterns in substantial corpora of textual and other data from across multiple domains. Despite their lack of specialist training, these models have demonstrated domain-specific capabilities that, it is inferred, could extend to the field of medicine. What is more, these assistants can be accessed via simple conversational interfaces, with users issuing prompts and receiving responses in plain language. This makes LLMs accessible in a way that, for the most part, earlier AI models were not.

The uses to which ChatGPT has been put in medicine include diagnostic tasks. Although varied in their results, applications in domains including ophthalmology,^3^ rheumatology,^4^ general internal medicine,^5^ and radiology^6^ have indicated that ChatGPT can tackle diagnostic challenges with a high degree of accuracy.

While research on the use of LLMs in epilepsy care is still in its infancy, the ability of these models to ‘understand’ conversational language suggests that they could be able to propose diagnoses on the basis of patients’ seizure descriptions. Unlike conditions which require complex physical examination or extensive tests, seizure disorders such as epilepsy and are largely diagnosed by a clinician listening to patients’ own account of their conditions.^7^ While this would seemingly make them a good fit for LLMs, they have, so far, only featured in a very small number of research applications.^8^

Even outside of epilepsy, research involving LLMs has tended to use vignettes and case reports rather than patients’ own accounts. Although such approaches are demonstrably effective, diagnosis based on a patient’s own words would suggest a more direct application of LLMs, closely aligned with the traditional doctor-patient interaction. This would be particularly useful early in the diagnostic pathway, before more highly specialised clinicians become involved.

With the present study, we aim to address both of these omissions by applying a LLM (GPT-4) to a diagnostic challenge commonly leading to mistreatment and delayed diagnoses^9^: the differentiation of epileptic and functional / dissociative seizures (FDS). This task continues to depend on the effective elicitation and expert interpretation of seizure descriptions from patients and witnesses.^10^ While symptom constellations are of proven diagnostic value,^11^ experts in the diagnosis of seizure disorders can extract additional diagnostic information by noticing *how* patients talk about seizure experiences, e.g. which aspects they highlight or volunteer, and how detailed seizure descriptions are.^12^ Indeed, studies involving recordings of patients speaking a range of different languages have demonstrated that linguists solely applying conversational criteria can differentiate between epileptic and functional seizure accounts with a high degree of accuracy.^13–17^ Building on these observations, Pevy et al.^18^ programmed an automatic machine learning model to extract linguistic features from transcripts of patients’ seizure descriptions. This model achieved a diagnostic accuracy of up to 81%.

The present study was intended to explore how well and reliably a non-specialist LLM-based classifier would tackle the differentiation of spoken descriptions of epileptic and functional seizures.

## 2. Methodology

### 2.1. Patients

Participants were recruited in two different ways. Patients over 16 years of age attending any seizure clinic at the Royal Hallamshire Hospital were sent information about this study with a reminder letter about two weeks prior to their face-to-face or telephone clinic appointment. Information about the study was also posted through various communication channels of the following charities supporting individuals with seizures: Epilepsy Action, FNDHope, FNDAction, Epilepsy Sparks, and the Shape network (supported by Epilepsy Research UK). Potential participants completed a ‘consent-to-contact’ form and were approached by a member of the research team. The study therefore represents a convenience sample.

The diagnosis of participants recruited via the Royal Hallamshire Hospital was confirmed using the medical records, whereas self-disclosure was used to ascertain the diagnoses of participants who were recruited externally. Participants indicated that they had been diagnosed either using video-EEG or through clinical assessment by a seizure specialist.

### 2.2. Data collection

Participants who had decided to take part in the research were directed to a website where, having provided informed consent, they could ‘interact’ with a ‘virtual agent’ (the talking head of a doctor presented on their computer screen). This agent was featured in eight videos embedded in a web application, which participants accessed via a secure login. Each video featured the agent asking one question (see Appendix 1 for a full list of questions), which participants would answer before clicking through to the next video. (Note that, partway through the study, we noticed that one of the virtual agent’s questions—but not the patient’s answer—was missing from a single transcript. We corrected the error and, given how minor it was, do not believe that it would have had an impact on the results. There was also a case where a single question-answer pair was missing from a transcript. This omission was present in the original data.)

Patients’ answers were audio-recorded as part of an earlier study into the automated collection and analysis of accounts of transient loss of consciousness (TLOC) via traditional machine learning methods.^19^ All participants had given permission for their answers to be used in further research. Note that, for ethical and practical reasons, we used only pseudonymised transcripts of the recordings for this study. We have included AI-generated examples of patients’ interactions with the digital doctor in Appendix 3. (See Pevy et al.^19^ for full details of the original data collection.)

Following removal of patients with syncope and those who had not consented to their recordings being used in future analyses, 41 transcripts from the earlier study were available for this study (16 of patients with epilepsy, 25 with FDS).

### 2.3. Model

The LLM we used in this study was one of OpenAI’s Generative Pretrained Transformers (GPTs), GPT-4. This is the same model that underlies some premium versions of ChatGPT. However, whereas ChatGPT has been further fine-tuned for conversation, we accessed GPT-4 directly using OpenAI’s Chat Completions application programming interface (API). This allowed us to write code to interact with the model in an automated way (see Procedure, below), an approach which is not possible through the standard ChatGPT interface. A further benefit of using the API is that it does not retain data for training purposes.

Accessing the model via the API also allowed us to adjust certain parameters not modifiable in ChatGPT. Although we left most parameters at their defaults, we did reduce the ‘temperature’ parameter to 0. Temperature is a parameter that controls the ‘creativity’ of a model’s response.^20^ A higher temperature setting generates more creative, less predictable answers, while a lower temperature setting generates less creative, more consistent answers. We set the temperature at its lowest possible value because the task that we were setting the model (classification) required constrained responses and because we wanted our results to be as replicable as possible. The API also has a ‘response_format’ parameter that allowed us to specify that the model should return its answers in a specific format (see below).

We used the programming language JavaScript (through the framework Node.js) to access the API. The specific GPT-4 model we used was gpt-4-turbo-2024-04-09, which was the latest model at the time we started conducting the research (see OpenAI^21^ for details of this model).

### 2.4. Conditions

We carried out four conditions to test GPT-4’s abilities. In the first condition, we wanted to test GPT-4’s ‘raw’ abilities—that is, its abilities based solely on its original training data. This meant that, in our prompt to the model, we described only the task that we wanted it to perform and the output that we wanted it to produce. This is often referred to as ‘zero-shot’ prompting^22^ because the model is being given no examples (i.e. ‘shots’) to guide its output. Because the zero-shot condition was our baseline, we also carried out several variations where we tweaked aspects of our prompt (e.g. using different terminology) and properties (e.g. raising the temperature setting) to see if they had any impact.

The remaining conditions were all one-shot or few-shot conditions—that is, they all involved us giving the model labelled examples of epilepsy and FDS accounts before asking it to diagnose the unlabelled example. In the one-shot condition, we gave it one example of epilepsy and one example of FDS. In the two-shot condition, we gave it two examples of each, and in the three-shot condition we gave it three examples of each.

These examples were randomly selected from among those cases that all three neurologists had correctly diagnosed. Providing shots is a well-established way of improving a model’s performance.^23^

To provide a benchmark for GPT-4’s performance, we had three experienced neurologists with particular epileptological expertise to classify the same cases using the same data. All of these neurologists had over 20 years of professional experience. As with GPT-4, they were given the transcripts in isolation, with no additional information.

### 2.5. Procedure

We prepared formatted programming objects for each of our cases. Each object featured an ‘id’ field (the IDs were reused from the earlier study), a ‘label’ field (‘epileptic’ or ‘FDS’), and a ‘transcript’ field (featuring the full transcript of the patient’s interaction with the digital doctor). All 41 objects were compiled in a formatted ‘array’ (i.e. a list) so that they could be manipulated and accessed programmatically.

We wrote a loop that would iterate over this array, extract the necessary information for each case, and combine it with our pre-written prompts in which we told GPT-4 what we wanted it to do for that condition. Table 1 shows the text of our prompts for the zero-shot and one-shot conditions, while Table 2 shows our variations on the zero-shot condition. (Note that the two-shot and three-shot prompts were identical to the one-shot prompt apart from the additional examples.)

**Table 1.**
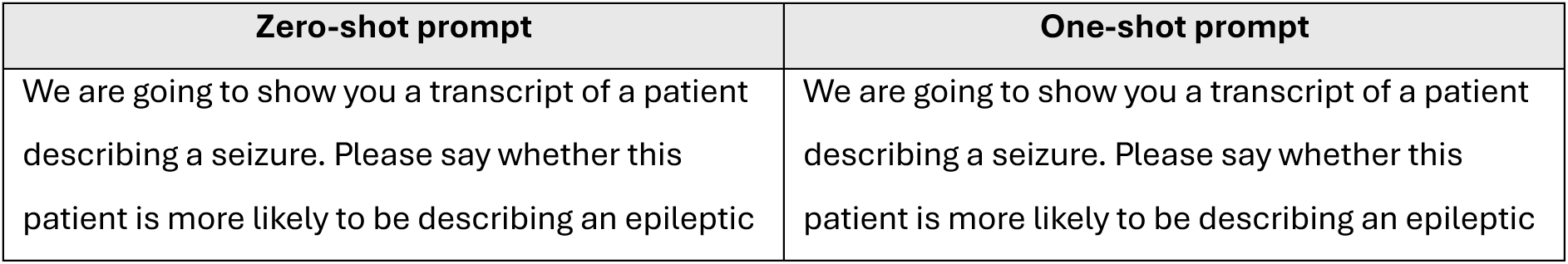
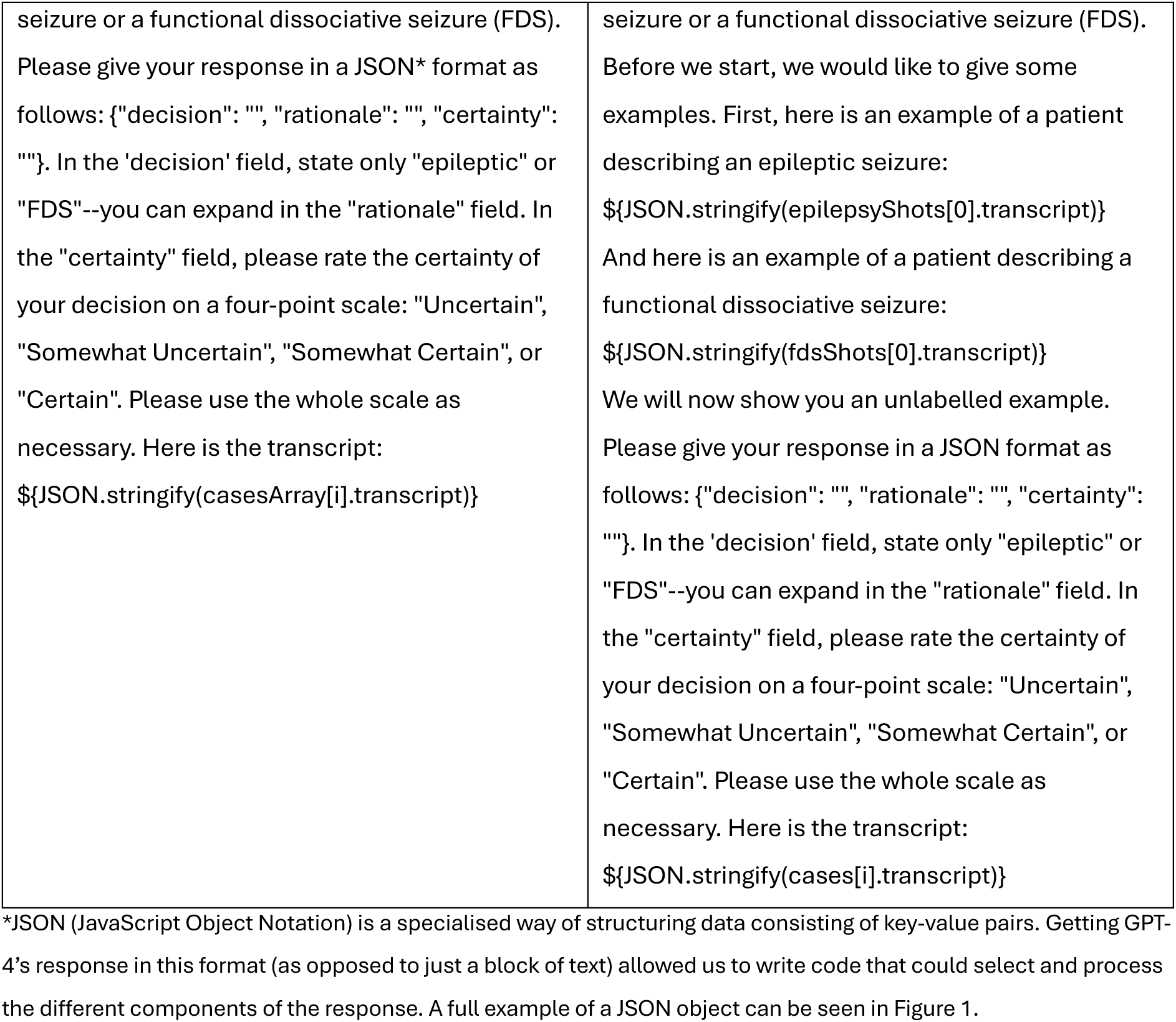
Prompts used for each condition.

| Zero-shot prompt | One-shot prompt |
| --- | --- |
| We are going to show you a transcript of a patient describing a seizure. Please say whether this patient is more likely to be describing an epileptic | We are going to show you a transcript of a patient describing a seizure. Please say whether this patient is more likely to be describing an epileptic |
| seizure or a functional dissociative seizure (FDS). Please give your response in a JSON* format as follows: {"decision": "", "rationale": "", "certainty": ""}. In the 'decision' field, state only "epileptic" or "FDS"--you can expand in the "rationale" field. In the "certainty" field, please rate the certainty of your decision on a four-point scale: "Uncertain", "Somewhat Uncertain", "Somewhat Certain", or "Certain". Please use the whole scale as necessary. Here is the transcript:<br>`{JSON.stringify(casesArray[i].transcript)}` | seizure or a functional dissociative seizure (FDS). Before we start, we would like to give some examples. First, here is an example of a patient describing an epileptic seizure:<br>`{JSON.stringify(epilepsyShots[0].transcript)}`<br>And here is an example of a patient describing a functional dissociative seizure:<br>`{JSON.stringify(fdsShots[0].transcript)}`<br>We will now show you an unlabelled example. Please give your response in a JSON format as follows: {"decision": "", "rationale": "", "certainty": ""}. In the 'decision' field, state only "epileptic" or "FDS"--you can expand in the "rationale" field. In the "certainty" field, please rate the certainty of your decision on a four-point scale: "Uncertain", "Somewhat Uncertain", "Somewhat Certain", or "Certain". Please use the whole scale as necessary. Here is the transcript:<br>`{JSON.stringify(cases[i].transcript)}` |

\*JSON (JavaScript Object Notation) is a specialised way of structuring data consisting of key-value pairs. Getting GPT-4's response in this format (as opposed to just a block of text) allowed us to write code that could select and process the different components of the response. A full example of a JSON object can be seen in Figure 1.

**Table 2.** Zero-shot variations.

| Variation | Reason | Modification |
| --- | --- | --- |
| Different terminology | FDS are referred to by different names throughout the literature that GPT-4 is likely to have been trained on. The use of one name over another could affect the model's performance. | Instead of referring to "functional dissociative seizures" in our prompt, we referred to "psychogenic non-epileptic seizures". |
| Diagnosis only | Asking a model to work through a problem can, in some circumstances, improve its performance.<sup>24</sup> This means that seeking a rationale from the model alongside the diagnosis could have affected the results. | We asked the model to provide only a diagnosis, leaving out the additional fields (rationale and certainty) that we had asked for in our standard zero-shot condition. |
| System message | A system message is a way of setting a model's behaviour.<sup>25</sup> | We set a custom system message that was tailored to the task at hand:<br>*You are SeizureClassifier, a helpful assistant in the field of neurology. Users will give you transcripts of patients describing seizure events. It is your job to look at the examples in detail and provide a diagnosis. Users may tell you to limit your diagnoses to a small number of options, which is acceptable and must be adhered to. When making diagnoses, you must not take the easy route. An example of the easy route would be "this patient said that they injured themselves, therefore, they must have epilepsy." Look in detail at what the patients are saying, consider both traditional and cutting-edge literature, and make a well-rounded, informed diagnosis commensurate with your level of expertise (which should be equivalent to that of an experienced neurologist).* |
| Temperature 0.5 | We set our baseline temperature at 0, for reasons described above. Temperature could affect the model's performance. | Temperature increased to 0.5. |
| Temperature 1 | As above. | Temperature increased to 1. |
| Temperature 1.25** | As above. | Temperature increased to 1.25. |

\* Although we tried raising the temperature to 1.5, it produced an output that was unstable and not formatted in the way that we had requested. We therefore set the cut-off at 1.25.

In the zero-shot condition, the loop simply extracted the transcript for each case because this was the only information we wanted to give to the model. In the one-, two-, and three-shot conditions, there was an additional step in which we extracted the training examples (shots) as well, which contained both the transcript and the label.

As can be seen in Table 2, we asked GPT-4 to return its answers in a particular format (JSON). Figure 1 gives an example of a response object. The ‘id’, ‘label’, and ‘transcript’ fields are all taken from the original object that we created when preparing the data. The ‘decision’, ‘rationale’, and ‘certainty’ fields have all been generated by GPT-4, while the ‘accurate’ field has automatically been set to false (a Boolean value) because the ‘label’ and ‘decision’ fields do not match.

**Figure 1.**
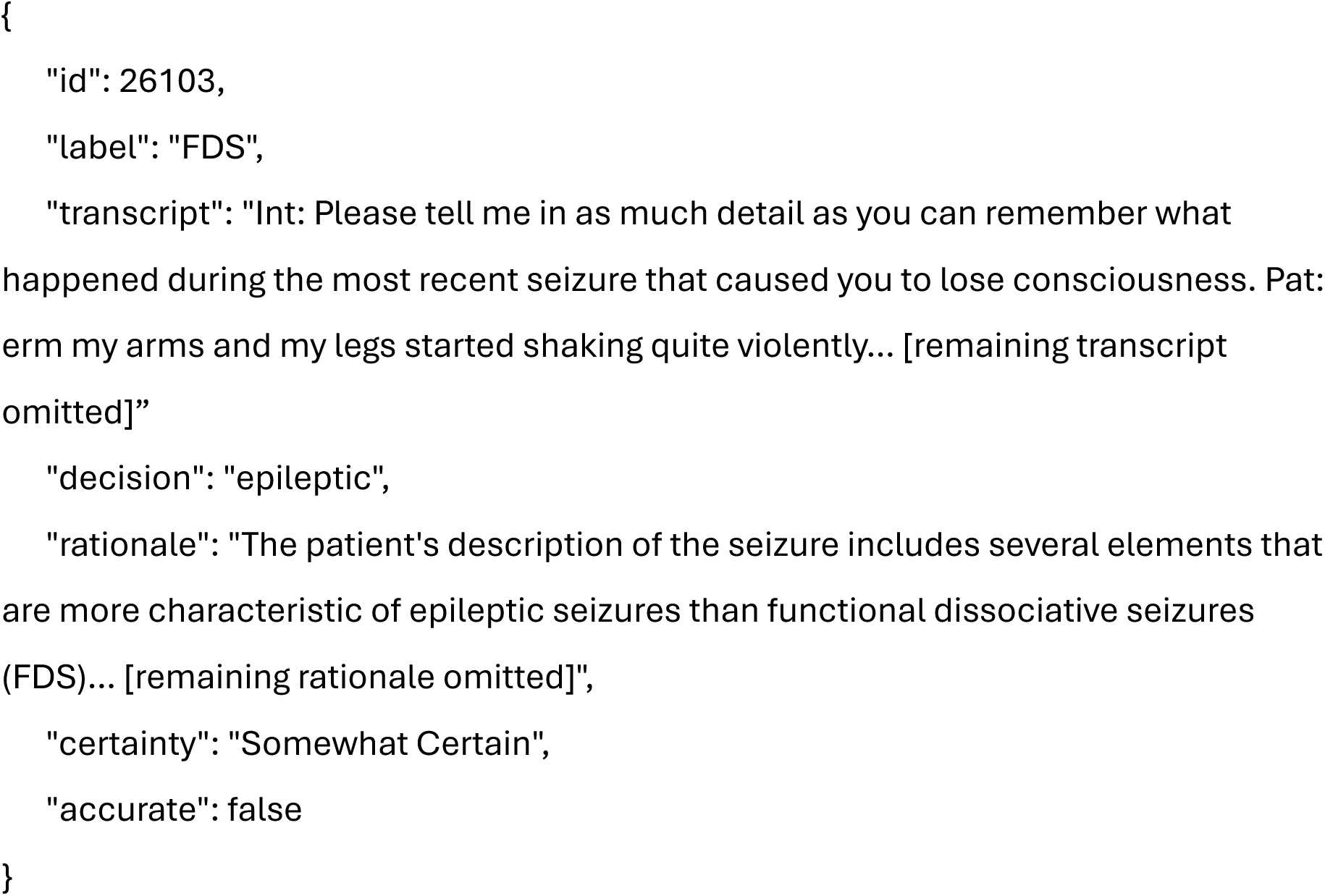
Example JSON response object

GPT models are non-deterministic—that is, the same input will not consistently yield the same output. Although lowering the temperature as we did makes them more deterministic, there is still room for some variability. For this reason, we had GPT-4 repeat each condition multiple times. For the zero-shot condition, we carried out three repetitions. For the few-shot conditions, we doubled this to six due the extra variability introduced by the randomly selected training examples.

### 2.6. Analysis

We analysed the data using R in R Studio. Most statistics were calculated using the Classification and Regression Training (caret)^26^ package’s confusionMatrix function. This function produces several statistics for testing the performance of classifiers. For reporting purposes, we have selected basic accuracy (i.e. the raw percentage of cases that were accurately classified), both overall and for epilepsy and FDS separately; balanced accuracy, which takes into account the proportion of accurate positives and accurate negatives; F1 score, a statistic which averages precision (i.e. how many diagnoses were correct) with recall (i.e. how many of the cases were correctly identified); a hypothesis test between overall accuracy and the no information rate (i.e. the accuracy that a classifier would achieve if it chose the majority case every time); and Cohen’s Kappa, to measure the strength of agreement between predicted and actual diagnoses. The confusionMatrix function also provides confidence intervals. We also used the IRR package^27^ to calculate inter-rater reliability (Fleiss’s Kappa) and the ggplot package^28^ to produce a visualisation.

#### Regulatory Approval

The Leicester South Ethics Committee reviewed and granted ethical permission for this research (REC reference: 20/EM/0106).

## 3. Results

The results for each condition are described below. Full results are shown in Table 3.

**Table 3.**
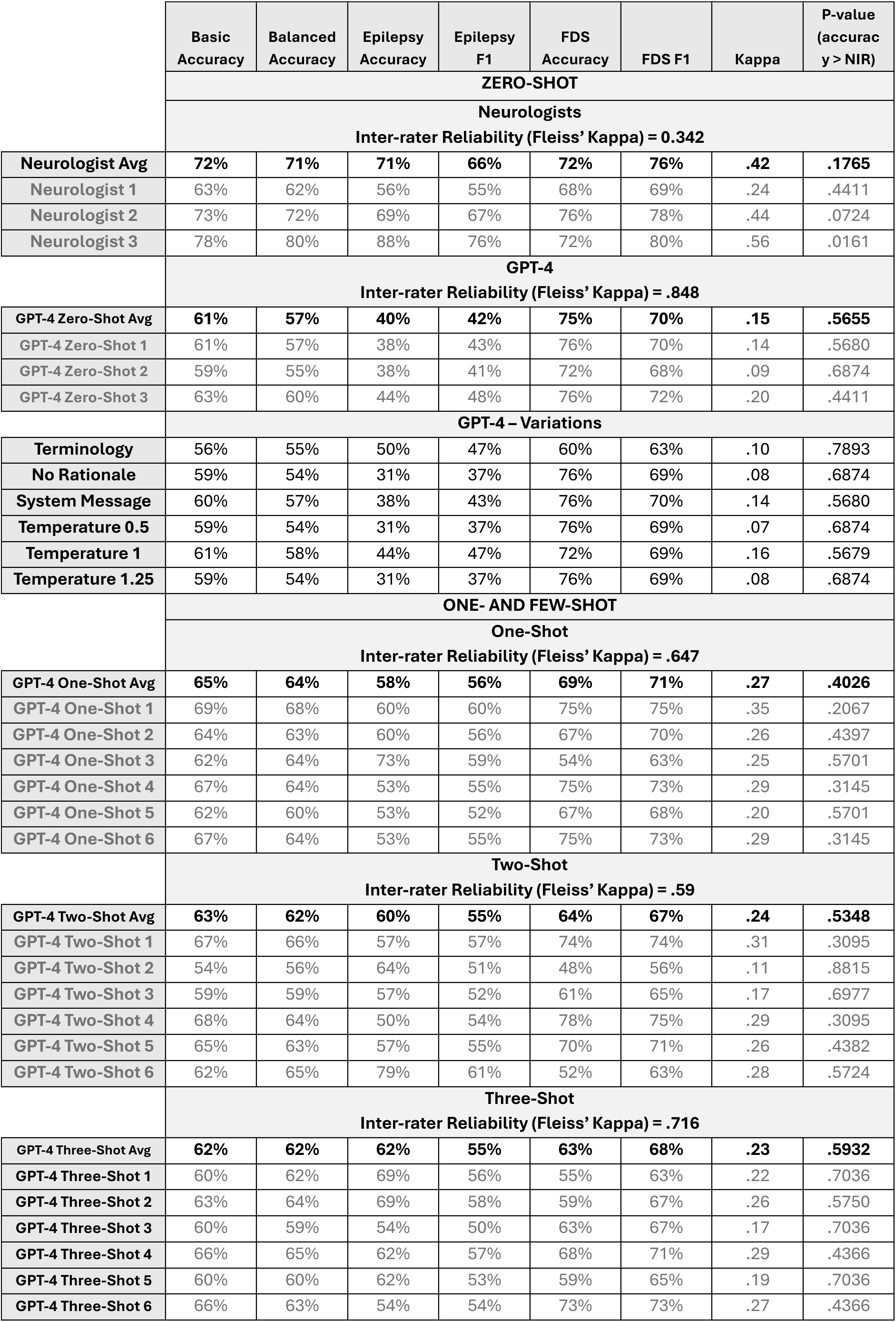
GPT-4 and neurologist performance.

### 3.1. Zero-Shot

GPT-4’s basic accuracy scores across the three repetitions of the task in this condition were 61% (95% CI: .445, .758), 59% (95% CI: .421, .737), and 63% (95% CI: .469, .779). Balanced accuracy (a value which takes into account the unequal epilepsy and FDS cases in our dataset) was 57%, 55%, and 60%. The hypothesis test between GPT-4’s accuracy and the ‘no information rate’ was non-significant in all three cases (p > .05). Cohen’s Kappa values were .1414, .0983, and .2044, indicating weak-to-no agreement between GPT-4’s diagnoses and the labels. Fleiss’ Kappa indicated strong agreement across the model’s different runs (.848).

All three neurologists performed better at the task than GPT-4, although there was noticeable variation between them, with basic accuracy scores of 63%, 73%, 78% and balanced accuracy scores of 62%, 72%, and 80%. Kappa scores were similarly varied, ranging from weak-to-no agreement for the first neurologist (.24) to moderate agreement for the second and third neurologists (.44 and .56, respectively).

Comparison between the accuracy and the NIR were non-significant for the first two neurologists (p > .05) but significant for the third neurologist (p < .05). Fleiss’ Kappa indicated moderate agreement among the neurologists (.342).

We calculated confusion matrices for GPT-4’s three repetitions and for the neurologists (Figure 2). These matrices show that GPT-4 was noticeably worse at identifying epilepsy cases (accuracy: 37%, 38%, and 44%) than FDS cases (76%, 72%, and 76%). This is also reflected in the F1 scores, which were 43%, 41%, and 48% for epilepsy and 70%, 68%, and 72% for FDS.

**Figure 2.**
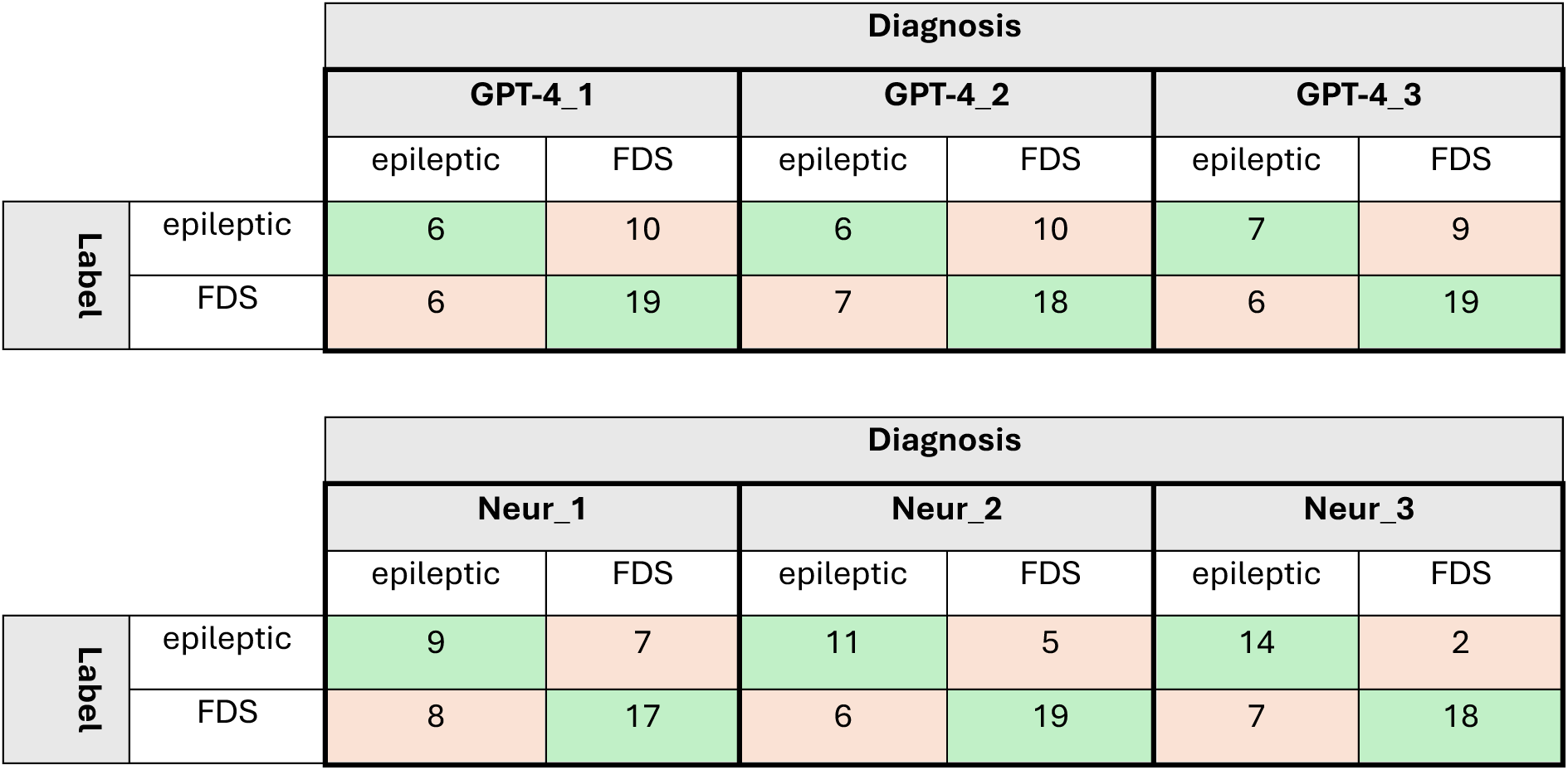
Confusion matrices for GPT-4 and neurologist performance in the zero-shot condition

The first and second neurologists were also worse at identifying epilepsy cases (accuracy: 56% and 69%, F1: 55% and 67%) than FDS cases (accuracy: 68% and 76%, F1: 69% and 78%), although the gaps were smaller and performance was always above 50%. There was no such disparity for the third neurologist, who was *more* accurate at identifying epilepsy cases (accuracy: 88%, F1: 76%) than FDS cases (accuracy: 72%, F1: 80%).

#### 3.1.1. Zero-Shot: Variations

We carried out several variations on the zero-shot condition for GPT-4. Only one of these (raising the temperature to 1) improved upon the averaged balanced accuracy of the baseline condition, and then only by a single percentage point. The remaining variations saw either equivalent or worse performance.

### 3.2. One- and Few-Shot Conditions

#### 3.2.1. One-Shot

GPT-4’s performance in the one-shot condition was noticeably improved from its performance in the zero-shot condition. Average basic accuracy across six repetitions was 65% (range: 62% - 69%), with a balanced accuracy of 64% (range: 60% - 68%). The locus of improvement was in the identification of epilepsy cases, which increased to an average accuracy of 58% (range: 53% - 73%) and an average F1 score of 56% (range: 52% - 60%). Average accuracy on the identification of FDS cases, however, dropped to 69% (range: 54% - 75%), with a similar drop in F1 score to 71% (range: 63% - 75%). The average Cohen’s Kappa was .27 (range: .20 - .35). Despite the improvement, GPT-4’s performance in this condition was worse than the group performance of the experienced neurologists. While noticeably weaker than the zero-shot condition, Fleiss’ Kappa still indicated strong agreement between the model’s different runs (.647).

#### 3.2.2. Two-Shot

GPT-4’s average performance in the two-shot condition was worse than its performance in the one-shot condition, though still better than its performance in the zero-shot condition. Average basic accuracy across six repetitions was 63% (range: 54% - 67%), with balanced accuracy of 62% (range: 56% - 66%), epilepsy accuracy of 60% (range: 50% - 79%), epilepsy F1 of 55% (range: 51% - 61%).), FDS accuracy of 64% (range: 48% - 78%), and FDS F1 of 67% (range: 56% - 75%). Average kappa was .24 (range: .11 - .31). Fleiss’ Kappa indicated moderate agreement between the model’s different runs (.59).

#### 3.2.3. Three-Shot

Average performance in the three-shot condition was almost identical to that of the two-shot condition, with basic, balanced, and epilepsy accuracies of 62% (range: 60% - 66%, 59% - 65%, 54% - 69%), FDS accuracy of 63% (range: 55% - 73%), and an average kappa score of .23 (range: .17 - .27). Epilepsy F1 was 55% (range: 50% - 58%) and FDS F1 was 68% (range: 63% - 73%). Fleiss’ Kappa indicated that agreement between the runs was stronger than either the one- or two-shot conditions (.716). Figure 3 shows GPT-4’s three-, two-, one-, and zero-shot average balanced accuracy compared to the neurologist average.

**Figure 3.**
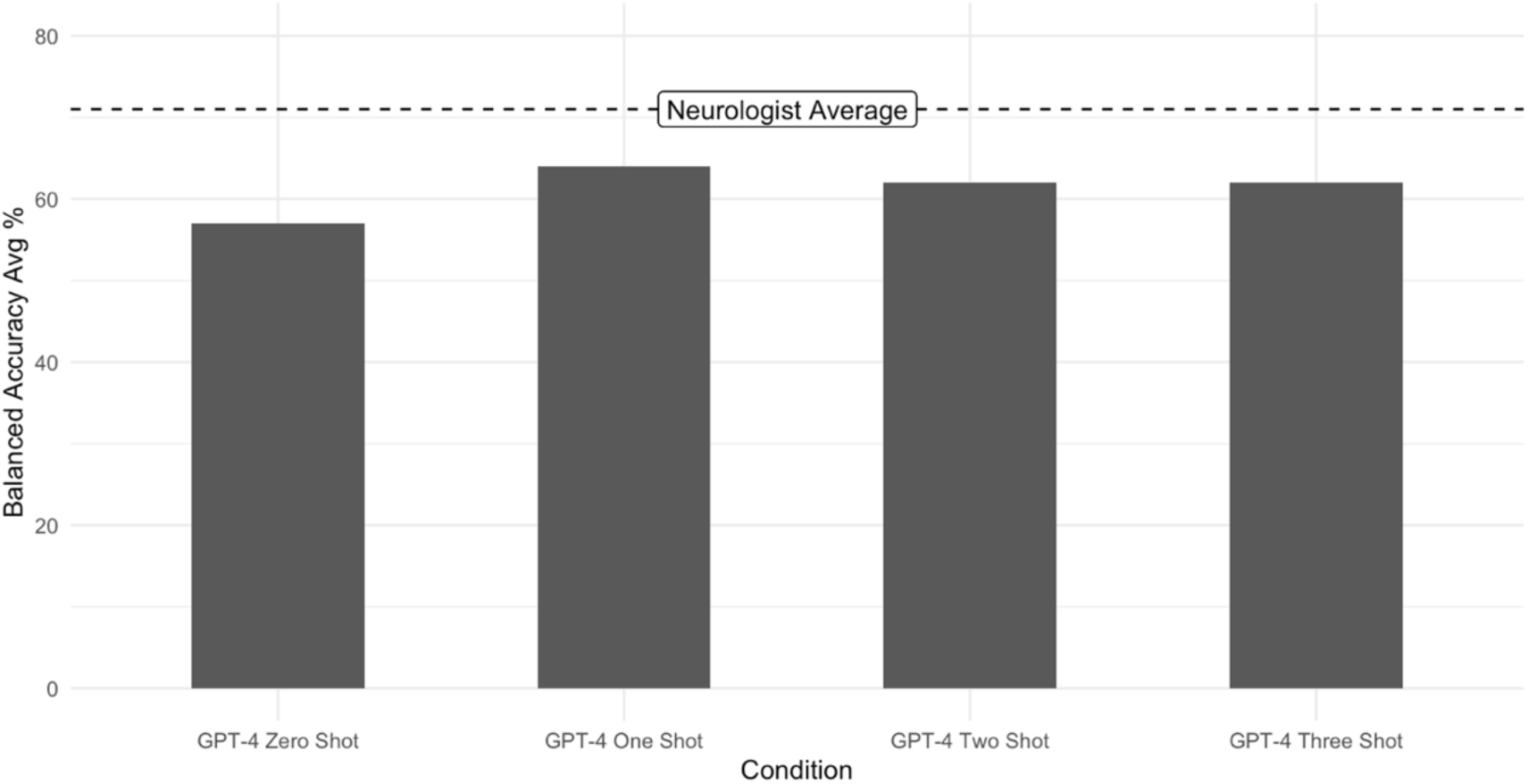
Average balanced accuracy by condition

## 4. Discussion

The aim of this study was to gauge the performance of a non-specialist LLM (GPT-4) on the notoriously difficult task of differentiating between epilepsy and FDS based on patients’ accounts.

We started by assessing GPT-4’s baseline (zero-shot) performance, which was hardly better than chance and substantially worse than the performance of experienced neurologists. GPT-4 was also noticeably less accurate at identifying epilepsy than FDS in this condition. While others have described how even small changes to prompt design can yield significant performance improvement on certain tasks,^22^ and that well-designed zero-shot prompts may outperform few-shot prompts in some cases,^29^ a range of prompt engineering approaches to improve the zero-shot approach failed to enhance its performance in the differential diagnosis of seizures. Although we cannot rule out the possibility that other changes to temperature settings, system message, or some combination thereof could have produced a striking improvement in the GPT-4’s diagnostic accuracy, this suggests that, rather than there being a flaw in our approach, GPT-4’s training data had not prepared it sufficiently well for the task that we had set for it. This may be because that training data was drawn from online corpora and would be more likely to contain written accounts of seizures (e.g. drawn from forums) rather than the transcribed spoken accounts that we used in the present study. It could also be that the training corpora contained low-quality information about criteria of diagnostic value.

The performance of GPT-4 improved noticeably in the subsequent test conditions, where we provided increasing numbers of randomly selected training examples in our prompts, particularly when identifying cases of epilepsy. While its performance remained worse than that of expert neurologists, the response of the model provides a glimpse of what LLMs may be able to achieve with more advanced approaches in the future (see below).

We note, however, that the performance improvement did not correlate with the number of examples that we provided. Indeed, the most substantial improvement came when we had given GPT-4 just one example of epilepsy and FDS, with the two- and three-shot conditions seeing worse (though not substantially worse) performance (see Figure 3). It is difficult to say whether this was a plateauing (i.e. any number of training examples will lead to basically the same improvement) or whether the additional shots were actively making the model’s performance worse. It has been suggested that providing examples does not always improve performance,^29^ and there is also evidence that LLMs can struggle to access information in the middle of longer inputs.^30^ The quality of the training examples (which were randomly selected) could also have had an impact, although we did exclusively train the model with examples which all neurologists had diagnosed correctly.

It may be that performance would start to increase again with more training examples. Even greater performance improvements could be achieved using more elaborate approaches than the ‘prompt engineering’ we have used in this study. This could mean, for example, fine-tuning a model with a larger number of cases from across a wider range of settings to tailor it to the specific task of classifying seizures. It could also mean implementing approaches such as retrieval augmented generation (RAG), where a model is able to draw upon deeper knowledge from more specialist sources before providing its answer.^31^ There is also the possibility of testing, or developing, other LLMs beyond GPT-4. These are all avenues for future research. Due to the rapidly developing nature of the field, there will also be ongoing improvements in LLMs themselves, both to the outputs that they produce and to their ability to ‘understand’ longer inputs.

Conceivably, the performance GPT-4’s could also be improved by combining its capabilities with the output of an automated diagnostic approach based on machine learning, with a model specifically trained to discover features of language previously found to help with the differentiation of accounts of epileptic and functional seizures. We note that the performance of our GPT-4 approach was noticeably worse than the best diagnostic accuracy observed by Pevy et al.^19^ using such an approach (which can likely to be attributed, again, to GPT-4’s lack of specialised training). While it is possible that, in view of the relatively small dataset, the model used in the previous study using pre-defined linguistic features would not generalise well to broader, more diverse samples of data, it is interesting that both approaches performed noticeably better when diagnosing FDS than epilepsy. Last but not least, the capability of an automated diagnostic approach involving GPT-4 might be improved by providing it with more detailed descriptions of seizures.

Notably, the ‘misdiagnosis’ rate of the clinicians who were asked to guess patients’ diagnoses on the basis of the relatively short transcripts of seizure descriptions without any additional information about the participants was much higher than one would expect from neurologists with particular expertise in the management of seizure disorders.^32^ This suggests that pivotal clinical information was missing from these particular seizures descriptions.

Our findings are aligned with a broad consensus emerging from research across various fields of medicine: that, while large language models show impressive performance (given their non-specialist training data) and great potential, they are currently lacking when it comes to more advanced medical reasoning. This consensus might best be captured by Hoomar et al.,^33^ who found that ChatGPT performed well on a first-year plastic surgery exam but significantly worse on more advanced exams.

Our study has a number of limitations: First and foremost, not all of our participants had ‘gold-standard diagnoses’, i.e. diagnoses proven by video-EEG. Participants recruited via online adverts were asked to confirm that their diagnoses had been made by an expert and they were recruited through advocacy organisations for their respective conditions. It is also the case that, by including participants without video-EEG proven diagnoses, our sample may have been more representative of the populations of people with epilepsy or FDS at large. Nonetheless, greater certainty of patients’ medical diagnoses would have bolstered our findings.

The nature of our data is also a potential flaw. We have already mentioned that the mode of data collection (with patients being asked questions in a linear, standardised way, with no opportunity for challenge, follow-up, or expansion built into the data collection platform) produced seizure descriptions which were much shorter and less detailed than accounts neurologists would typically settle for in a seizure clinic. Future research might explore LLMs’ diagnostic performance when presented with more naturalistic, interactional patient accounts, with a human neurologist and greater detail.

We conclude that, in view of these limitations, and despite the limited diagnostic capabilities of GPT-4 in the zero-shot condition, the responsiveness of this LLM to limited training others a tantalising glimpse of how LLMs may support diagnostic decision making, especially in settings where experts are not available.

## Data Availability

Data are not available for sharing.

## Acknowledgements

This research project was funded by Epilepsy Research UK (grant number 160296-1).

## Appendix 1: Questions asked by digital doctor*

1. Please tell me in as much detail as you can remember what happened during the most recent seizure that caused you to lose consciousness.
2. What were you doing when the seizure started and how were you feeling?
3. Do you think there was a trigger for the seizure?
4. What was the first sign of the seizure?
5. How did you feel during the seizure?
6. How did the seizure end?
7. How did you feel after the seizure?
8. Did you injure yourself during the seizure?

*Note that in some transcripts, the word ‘attack’ was used instead of ‘seizure’ in the questions. This change was made during original study when it was realised that ‘seizure’ would not cover cases of syncope.

## Appendix 2: Full results

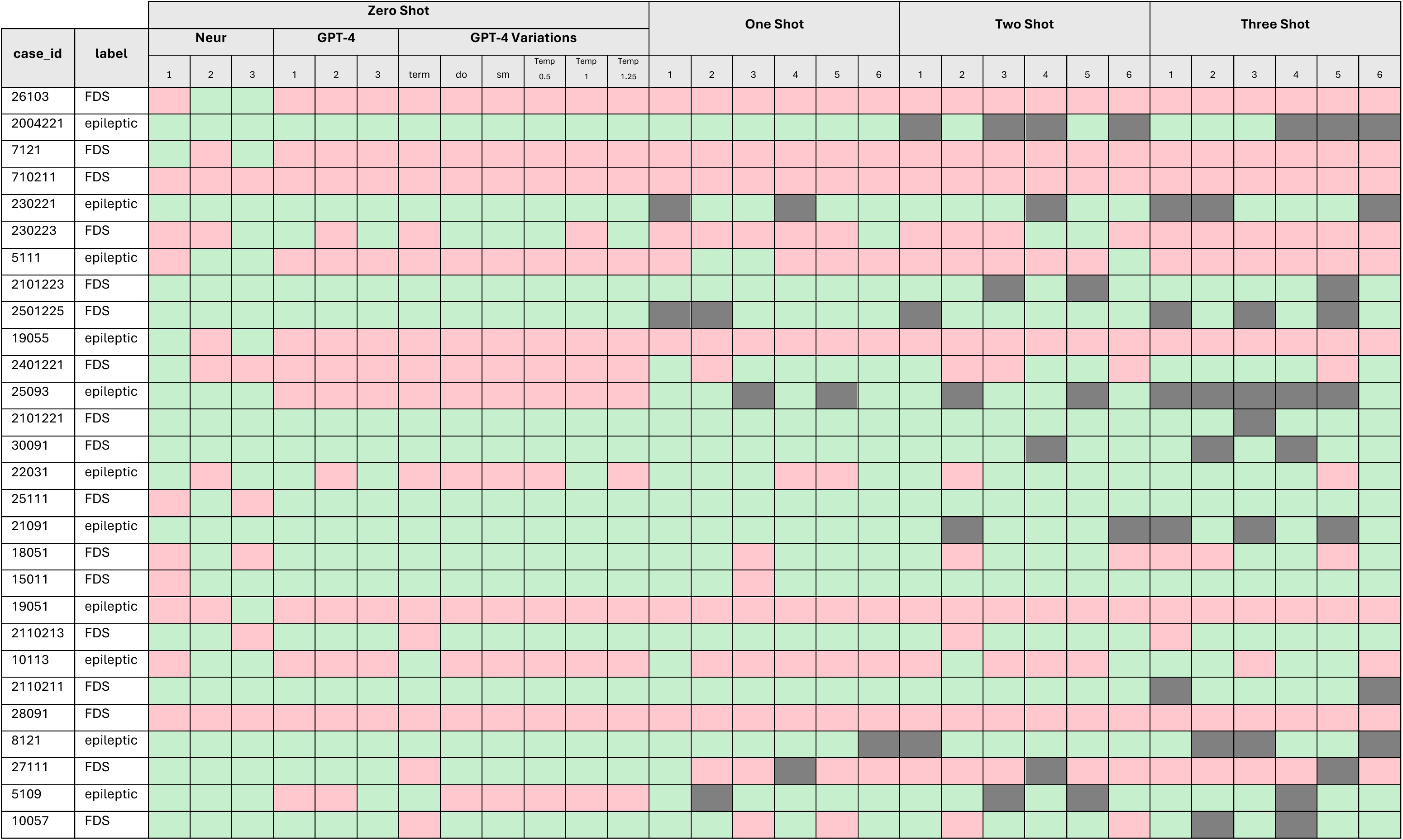
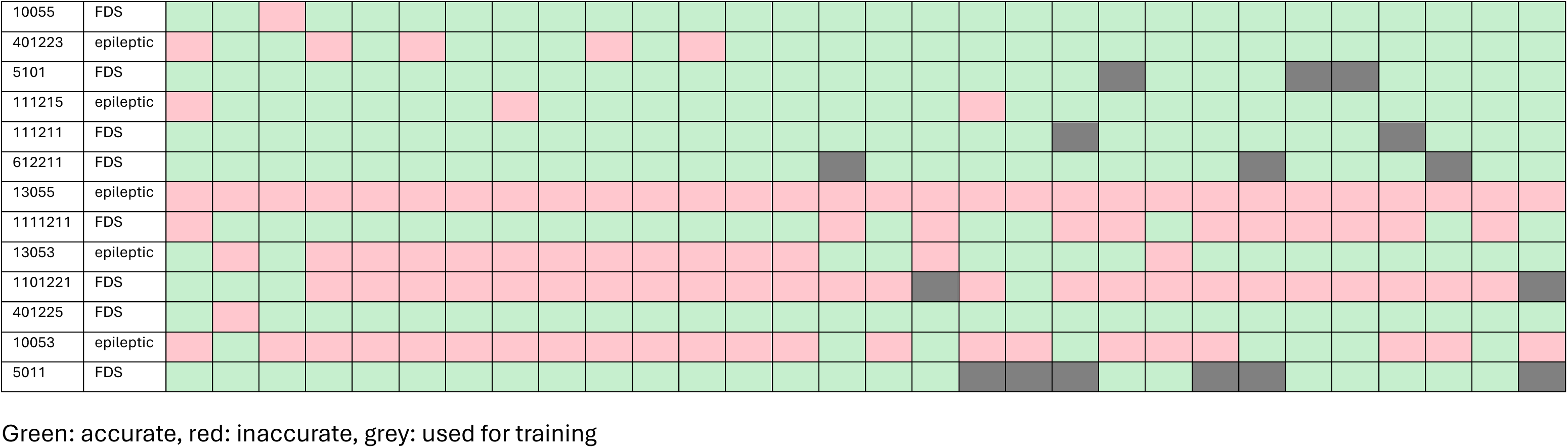

## Appendix 3: AI-generated examples of epilepsy and FDS digital doctor interactions

For ethical reasons, we are not able to include real examples of complete interactions in this article. However, we have given GPT-4 several examples for each condition and asked it to generate two fake interactions based on those. We used the same version of GPT-4 that we used for the main study (gpt-4-turbo-2024-04-09).

### 1. AI-generated example of FDS description

**Int:** Please tell me in as much detail as you can remember what happened during the most recent seizure that caused you to lose consciousness?

**Pat:** (2 seconds) Um, yeah, the last time I think it was about (1 second) two months ago. I was at this garden party, and um, I just started to feel really oA, like you know out of nowhere. First, I felt this wave, kind of (1 second) like you get chills? Yeah, chills but my head was just spinning. And the lights and the sounds started getting really (1 second) um annoying I guess, overwhelming. (3 seconds) And then I don’t remember much, just like, snippets, flashes of people, sounds, really blurred.

**Int:** What were you doing when the seizure started and how were you feeling?

**Pat:** I was just chatting, standing around with some friends, feeling pretty good until then, I hadn’t noticed anything odd before that moment (1 second) um suddenly just this switch, and I was down. Didn’t even feel it coming apart from that initial weird chill.

**Int:** Do you think there was a trigger for the seizure?

**Pat:** (2 seconds) Um, maybe? I don’t know. It could have been the heat? It was quite (1 second) hot, and the party was crowded, a lot going on, um maybe too much for me at that moment.

**Int:** What was the first sign of the seizure?

**Pat:** That chill, like a sudden (1 second) wave going through me, accompanied by a kind of faintness. And then things just spiralled out from there, it was (1 second) really fast, to be honest.

**Int:** How did you feel during the seizure?

**Pat:** I don’t remember feeling much during the actual moment, it was like I was not quite there, you know? Just bits and pieces till I came around.

**Int:** How did the seizure end?

**Pat:** (2 seconds) Uh, I woke up on a couch inside, so someone must have moved me away from the crowd. Someone said I was out for just a couple minutes, and they had put a cushion under my head.

**Int:** How did you feel after the seizure?

**Pat:** Just disoriented (1 second) um, and really tired actually. Everyone was asking if I was okay, but it took me a bit to even understand how I got inside the house. A bit shameful, but yeah, just part of (1 second) dealing with this.

**Int:** Did you injure yourself during the seizure?

**Pat:** Luckily, no, not this time. People around me reacted fast so, (1 second) yeah, managed not to hurt myself falling or anything like that. But it’s always a worry, you know?

### 2. AI-generated example of epilepsy interaction

**Int:** Please tell me in as much detail as you can remember what happened during the most recent seizure that caused you to lose consciousness.

**Pat:** (2 seconds) Um, I was at the grocery store, right, picking out vegetables (1 second) and, and suddenly (0.5) I was on the ground. I-I don’t precisely know when I blacked out, but I was definitely out. (1 second) Um, they said I was shaking and all.

**Int:** What were you doing when the seizure started and how were you feeling?

**Pat:** Let’s see um, I was checking the tomatoes, you know, squeezing them (laughs) and, well, I felt this wave of, um, dizziness and sort of a, uh, heaviness in my chest (1 second) right before it happened.

**Int:** Do you think there was a trigger for the seizure?

**Pat:** Not sure, um, it might’ve been the lights or maybe I was just tired or hungry, um, didn’t really think much about it till (1 second) it hit me.

**Int:** What was the first sign of the seizure?

**Pat:** Uh, definitely the dizziness (0.5) then, like, this buzzing noise in my ears, kinda like when your ears ring but much louder, that kind of startled me, um (1 second), then blank, nothing until I woke up on the floor.

**Int:** How did you feel during the seizure?

**Pat:** Uh, I, I can’t remember anything from the actual moment. It’s just blackout, no memories (1 second), nothing until I came around and saw people around me.

**Int:** How did the seizure end?

**Pat:** I guess when I woke up, people were hovering over me, someone had called an ambulance, and slowly (1 second) things started making a bit of sense, but I was fuzzy.

**Int:** How did you feel after the seizure?

**Pat:** Exhausted, confused and a bit scared really, not knowing how you got to the floor (0.5) it’s unsettling (1 second). And a bit embarrassed you know, with everyone staring.

**Int:** Did you injure yourself during the seizure?

**Pat:** Fortunately, no, um, someone caught me before I fell completely, so besides being a bit shaken up, no bruises or bumps.

